# Prevalence of anxiety disorder among workers with cancer in Amman, Jordan

**DOI:** 10.1101/2020.04.08.20058032

**Authors:** Abdel Rahman Aref Ali Abu Shreea, Lee Khuan, Sharifah Norkhadijah Syed Ismail, Nasrudin Subhi, Sobuh (Moh’d Sobhi) Ahmad Abu-Shanab, Zaher Juma’ Raja Mashrqi, Irniza Rasdi

## Abstract

**Introduction:** Cancer is now being recognised as a long term conditions due to advances in treatments that increase the survival rate of patients with cancer to as long as 10 years from the time of the disease. Anxiety is among the commonly discovered psychiatric illness in patients with cancer and is often neglected. Approximately 10% of patients with cancer are affected with anxiety worldwide. Hence, this study was aimed to determine the prevalence of anxiety disorder and its associated factors among workers with cancer in Jordanian population.

**Methods:** A cross-sectional study was conducted at the King Husain Cancer Centre (KHCC) in Amman, Jordan. Proportional sampling technique was used to obtain the sample population of 355 workers with cancer. Data were collected through self-administered Generalized Anxiety Disorder (GAD-7) questionnaire and analyses were carried out using SPSS version 25.

**Results:** Response rate of 100% were obtained from the participants. Prevalence of anxiety disorder among workers with cancer was recorded at 20.8% with male (23.1%) having the higher prevalence rate than female (17.3%) workers with cancers. A significant difference in anxiety between marital status (*p*=0.025), types of cancer (*p*=0.001), treatment types (*p*=0.024) were observed. A multiple regression was run to predict anxiety disorder from marital status, type of cancer and treatment types. These variables statistically significantly predicted anxiety disorder [F(3, 351) = 8.117, *p* < .001, R_2_ = 0.225].

**Conclusion:** There is high prevalent of anxiety disorder among workers with cancer in Jordan. Predictors of anxiety among workers with cancer were also identified in this report.

## Introduction

Cancer is the second leading cause of death around the world, and is resulting to an estimated 9.6 million deaths in 2018 especially in low-and middle income countries [1]. The most common cancer risk factor is use of tobacco and is responsible for approximately 22% of cancer deaths[2]. Hepatitis and human papilloma virus (HPV) that are known as cancer causing infections are responsible for up to 25% of cancer cases in low-and middle-income countries[3]. The economic impact of cancer is significantly increasing. The total annual economic cost of cancer in 2010 was estimated at approximately US$ 1.16 trillion [4]. In Jordan, cancer is the second leading cause for death after heart related diseases (Ashraf, & Ahmad, 2015). Additionally, it is observed that cancer morbidity and mortality is to be increased as young people age. Focusing over the aetiology of cancer, it is observed that tobacco is one of the major causes for cancer in Jordanians (Ahmad, 2015). High prevalence of smoking is linked with high incidents of lung cancer that is most common type of cancer in Jordan. Other common types of cancer observed in Jordan include colon and bladder cancer. In the year 2010, only 41 cases of cancer were registered. However, as compared to past decades, it is realized that new cancer cases diagnosis has increased among Jordanians (MOH, 2017).

Anxiety disorder may negatively affect patients with cancer within the duration of their diagnosis. Jordanian cancer patients are confronted with worries and uncertainties related to the effects of cancer on their life. The fear of pain, progression of the cancer, death, spiritual questioning, and guilt are high in these patients[5]. Anxiety tends to increase as a result of cancer treatment trajectory[6], and is identified as the most reported symptom in patients with cancer. From the initial diagnosis to initiation of treatment, cancer recurrence, and the failure of treatment along with perception of dying increases the overall stress and anxiety in the patients[7]. Hence, anxiety is often termed as a response to cancer diagnosis, which is the normal behaviour towards the initial shock, disbelief, and emotional distress[8]. [9] indicated that the cancer patients continuously worried and fear about their future as well as cancer recurrence. Anxiety has an impact on physiological and psychological health and well-being of an individual, however, the symptoms might differ from individual to individual. Apart from physical pain, and emotional distress, anxiety and depression are common factors affecting cancer patients worldwide. Thus, it is very important to address this problem in especially workers suffering silently with cancer in Jordanian population.

Literatures are limited about the prevalent symptoms of anxiety in Jordanian population. However, several factors have been reported to be linked with anxiety in patients with cancer and these includes social support, sociodemographic and socio-economics variables as well as functional status[10–12]. Factors associated with anxiety in patients with cancer need to be thoroughly investigated to improve their quality of life[10, 13] as well as their health outcome. A number of studies were carried out on anxiety associated with cancer patients, but none has been carried out on workers with cancer particularly among Jordanian population. Thus, this study aimed to determine the anxiety disorder level and its associated factors, explore the sociodemographic, and clinical features of cancer among workers with cancer in Jordanian population.

## Materials and methods

### Study Design

In this work, a cross sectional study was carried out in King Husain Cancer Centre (KHCC) from October 2019 to January 2020. [14] stated that cross sectional research design is based on observational research design. Here, the researcher examined the outcomes as well as exposure for research participant at the same time. Participants are selected on the basis of a defined inclusion and exclusion criteria.

## Selection criteria

The eligibility criteria in this research were participants must be the citizen of Jordan and obtained the cancer treatment from King Husain Cancer Centre (KHCC). The patient must be the employee of any organization in Jordan. Whilst workers with cancer disease who are not Jordanian were excluded from the study. Patients who were not attending KHCC for their cancer treatment. Patients with cancer who are not unemployed were also excluded from the study. In addition, patient below 18 years age group and those that are not working in any organization were excluded.

## Determination of sample size

Sample size calculation was made using [15] formula for calculating samples size as below and the highest prevalence of the attribute (30.3%) was obtained and used to determine the number of sample required for this study [16].

Sample size (n) = Z1 -α/2 Pq /d^2^

= Z1^2^ -α/2 P(1-P) /d^2^

Where, Z1-α/2 = standard error when α = 0.05 (95% Confidence Interval) = 1.96

q = 1-P

P = prevalence of the attribute = (30.3%) [16]

d = Acceptable difference using 5% (0.05)

N = number of sample size

A total of 325 sample or respondents was required and after adding 10% (32.5) attrition rate, the sample size was = 357.5 ∼ 358. Therefore, we enrolled 358 workers with cancers in this study.

## Sampling method

The samples are to be drawn from accessible population working class of cancer patients in King Husain Cancer Centre (KHCC). Each of the patients considered independent unit and proportional sampling technique was used to obtain the sample population.

## Research instrument

Data were collected using a self-administered online questionnaire. The questionnaire was divided into five different sections with each section having questions related to its title. In summary, the questionnaire contains socio-demographic section, cancer disease information, workplace support system, work related issues, and the Generalized Anxiety Disorder section scale (GAD-7) that is adopted since it is one of the most widely used diagnostic self-report scales for screening, diagnosis and severity assessment of anxiety disorder. This scale comprised of seven questions and it is rated based on the whole scale score that was range from 0 to 21 and cut-off scores for mild, moderate and severe anxiety symptoms are 5, 10 and 15 respectively [17]. This section contains 7 items questions and the rating scale was based on 7 Likert system of measurement ranging from 1 = strongly disagree to 7 = strongly agree. A pilot study was conducted using Cronbach’s alpha. The GAD-7, WRI and WSS questionnaires were at an alpha (α) = 0.80, α = 0.843, and α = 0.913 respectively.

## Statistical Analysis

Data analysis was carried out using IBM Statistical Package for Social Sciences (SPSS) version 25. Descriptive statistics such as frequencies, percentages and exploratory analysis were used in normalizing the data and expressed as Mean±SD. One way ANOVA and Mann-Whitney U test were used in analysing normally and not normally distributed data. Multilinear regression was used to analyse predictors for anxiety disorder. The internal reliability of information is examined by use of Pearson correlation and Cronbach alpha test. The alpha value higher than 0.7 is perceived as satisfactory and internal consistency also provides the estimation of test-retest reliability. The significance of such a correlation was tested from *t*-test and *p*-value < 0.05 was considered statistically significant.

## Ethical consideration

Ethical approval was obtained from the University Ethics Committee for Research involving Human Subjects, Faculty of Medicine and Health Sciences (FMHS), University Putra Malaysia (UPM) with reference number: UPM/FPSK/JKPP/A0426. Permission and approval were also obtained from Institutional Review Board (IRB) of King Husain Cancer Centre (KHCC) with reference number: 19 KHCC 112. Participants were given a written informed consent with an appropriate ethical consideration regarding the information about the study, the right of withdrawal, and protect their confidentiality regarding their identity and the information that they did not wish to disclose.

## Results

### Sociodemographic characteristics of the respondents

A total of 355 workers with cancer were approached in the KHCC during the period of data collection. The response rate of the study were 100%. Table 1 shows the socio-demographic characteristics of the respondents. The mean age of the respondents was 42.3 years (95% Cl = 41.3, 43.4). There was significant difference (t = 1.921, df = 353, *p* = 0.05) between the mean age of male (43.1 years, 95% Cl = 41.7, 44.5) and that of female (41.0 years, 95% Cl = 39.5, 42.5) respondents in this study. The majority of the respondents were married (78.9%) and only 14.4% were single while 3.1% were divorced. Majority of them had a Bachelor (57.2%) with 12.4% had a high school qualification. In terms of their job role, business was the highest (29.9%), then followed teacher (20.6%) and health professional (20.0%) with only 18.0% were civilian workers while the least were drivers (2.3%).

**Table 1:** Distribution of the respondents according to socio-demographic characteristics (n = 355)

| **Variable(s)** | **Number** | **Percentage** |
| --- | --- | --- |
|  |  | **(%)** |
| **Age group (years)** |  |  |
| 19-30 | 50 | 14.1 |
| 31-40 | 111 | 31.3 |
| 41-50 | 117 | 33.0 |
| 51-60 | 77 | 21.7 |
| **Total** | **355** | **100.0** |
| **Gender** |  |  |
| Male | 216 | 60.8 |
| Female | 139 | 39.2 |
| **Total** | **355** | **100.0** |
| **Marital status** |  |  |
| Single | 51 | 14.4 |
| Separated | 3 | 0.8 |
| Divorced | 11 | 3.1 |
| Windowed | 10 | 2.8 |
| Married | 280 | 78.9 |
| **Total** | **355** | **100.0** |
| **Educational level** |  |  |
| Primary school | 16 | 4.5 |
| Intermediate school | 20 | 5.6 |
| High school | 44 | 12.4 |
| Two years college | 36 | 10.1 |
| Bachelor | 203 | 57.2 |
| Postgraduate education | 36 | 10.1 |
| **Total** | **355** | **100.0** |
| **Job role** |  |  |
| Business | 106 | 29.9 |
| Civil servant | 64 | 18.0 |
| Driver | 8 | 2.3 |
| Engineer | 33 | 9.3 |
| Health professional | 71 | 20.0 |
| Teacher | 73 | 20.6 |
| **Total** | **355** | **100.0** |

### Clinical characteristics of the respondents

Table 2 shows the clinical characteristics of workers with cancer. Majority of them (52.1%) were of stage II cancer stage and then followed by those in stage III (23.9%). The remaining patients were of stage I (12.7%) and stage IV (11.3%) respectively. In terms of treatment types received by the patients, majority received chemotherapy (91.5%) while the remaining have had surgery (3.7%), and received immunotherapy (3.4%) and radiotherapy (1.4%). When categorised based on cancer types, majority were of breast cancer (16.9%), followed by lung (13.2%), lymphoma (13.0%), colorectal (11.0%) and others accounting 10.7% and the least was cervical (1.7%). As for the duration of diagnosis of the disease (cancer), majority of the respondents reported having 1-5 months (49.6%), 251 months and above (40.8%), 6-50 months duration (6.2%) and the least were those who were diagnose within 201-250 months (0.3%).

**Table 2:** Clinical characteristics of the respondents (n = 355)

| **Characteristics** | **Number** | **Percentage** |
| --- | --- | --- |
|  |  | **(%)** |
| **Cancer stage** |  |  |
| Stage I | 45 | 12.7 |
| Stage II | 185 | 52.1 |
| Stage III | 85 | 23.9 |
| Stage IV | 40 | 11.3 |
| **Total** | **355** | **100.0** |
| **Treatment types** |  |  |
| Surgery | 13 | 3.7 |
| Chemotherapy | 325 | 91.5 |
| Radiotherapy | 5 | 1.4 |
| Immunotherapy | 12 | 3.4 |
| **Total** | **355** | **100.0** |
| **Type of cancer** |  |  |
| Bladder | 4 | 1.1 |
| Bone | 25 | 7.0 |
| Brain | 13 | 3.7 |
| Breast | 60 | 16.9 |
| Cervical | 6 | 1.7 |
| Leukaemia | 28 | 7.9 |
| Lung | 47 | 13.2 |
| Ovarian | 8 | 2.3 |
| Colorectal | 39 | 11.0 |
| Lymphoma | 46 | 13.0 |
| Pancreatic | 8 | 2.3 |
| Stomach | 12 | 3.4 |
| Testicular | 14 | 3.9 |
| Thyroid | 7 | 2.0 |
| Others | 38 | 10.7 |
| **Total** | **355** | **100.0** |
| **Duration of diagnosis** |  |  |
| 1- 5 | 176 | 49.6 |
| 6-50 | 22 | 6.2 |
| 51-100 | 8 | 2.3 |
| 101-150 | 3 | 0.8 |
| 201-250 | 1 | 0.3 |
| 251 and above | 145 | 40.8 |
| **Total** | **355** | **100.00** |

### Comparison of age based on mean rank according cancer stage

Table 3 shows the comparison of age based on the mean rank according to the cancer stage. The mean rank of ALL respondents (178.0) was lower compared with cancer stage III and stage IV but higher compared with cancer stage I and stage II. To determine the difference of mean rank age among cancer stages, one-way ANOVA was carried out. The analysis results showed that there was significant difference among the three stages of cancer in terms of mean rank age *F* (109, 171) = 9.892, *p* = 0.001). Patients with stage I and stage II cancer (140.2 vs 168.1) were younger compared with patients with stage III and IV cancer (186.2 vs 249.1) (*p* = 0.001). In contrast, patients with stage II were older than patients with stage I cancer (*p* = 0.001).

**Table 3:**
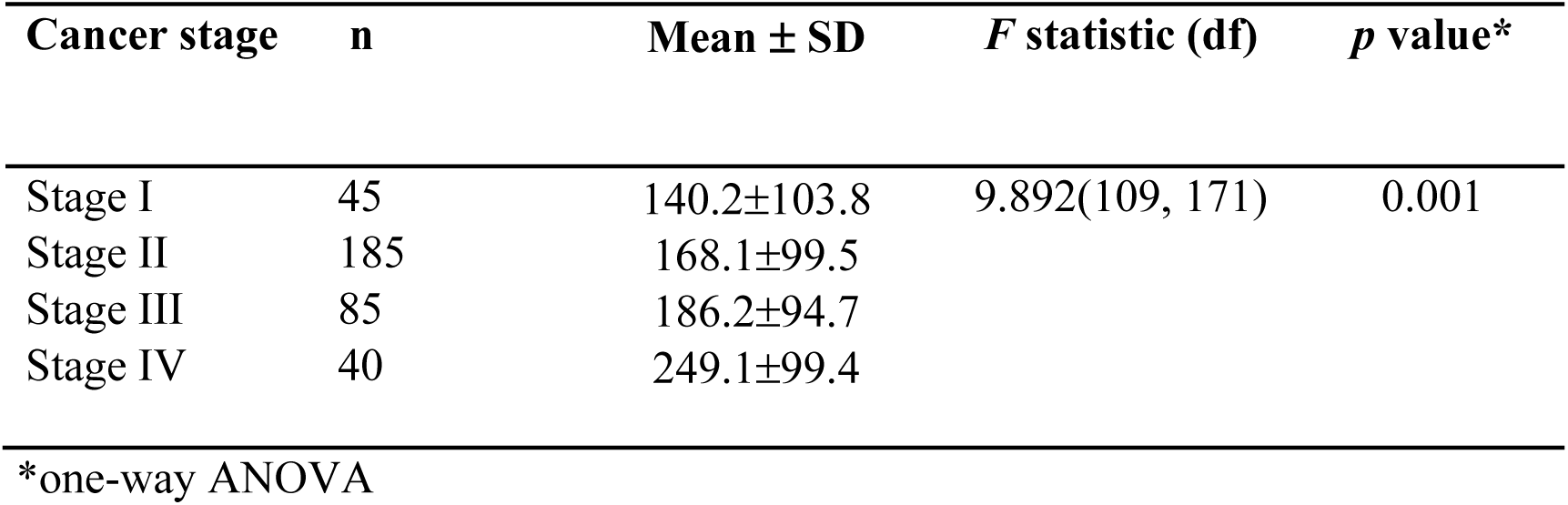
Comparison of age based on mean rank according cancer stage

### Prevalence of anxiety disorder and its severity

Table 4 shows the prevalence of anxiety disorders among workers with cancer. From the total of 335 respondents, the overall prevalence of the anxiety disorders was 20.8%. From 74 number of severity, 1.4% were found to have mild anxiety disorder, 11.5% were moderately severe and 7.9% were found to be severe. Severity among these patients is shown in Table 4 below.

**Table 4:** Severity of symptoms of anxiety disorder among workers with cancer (n=355)

| **Anxiety (GAD7)** | **Frequency** | **Percentage** |
| --- | --- | --- |
| **Mild** | 5 | 1.4% |
| **Moderate** | 41 | 11.5% |
| **Severe** | 28 | 7.9% |
| **Total** | 74 | 20.8% |

Moreover, this study found higher prevalence rate of anxiety in male (23.1%) compared to female (17.3%) workers with cancers. Among these respondents, there was higher preponderance of those whose marital status were Separated (33.3%), followed by Widowed (30.0%), Divorced (27.3%), Single (17.0%) and Married (20.7%). There was also a high proportion of anxiety disorder among workers with cancer whose educational level were intermediate school (35.0%), high school (27.3%), two year college (22.2%), and bachelor (19.7%). Similarly, anxiety disorder was found to be higher among Engineers (30.3%), and Business (25.5%) then followed by Driver (25.0%), Civil servant (20.3%) and Teachers (16.4%). Differences in mean of anxiety disorder were also determined, and there was a significant difference in anxiety between marital status (*p*<0.05), types of cancer (p<0.05), treatment types (p<0.05). However, there was no significant difference in anxiety disorder in educational level (p>0.05), job role (p>0.05), cancer stage (p>0.05) and gender (p>0.05) (Table 5).

**Table 5:**
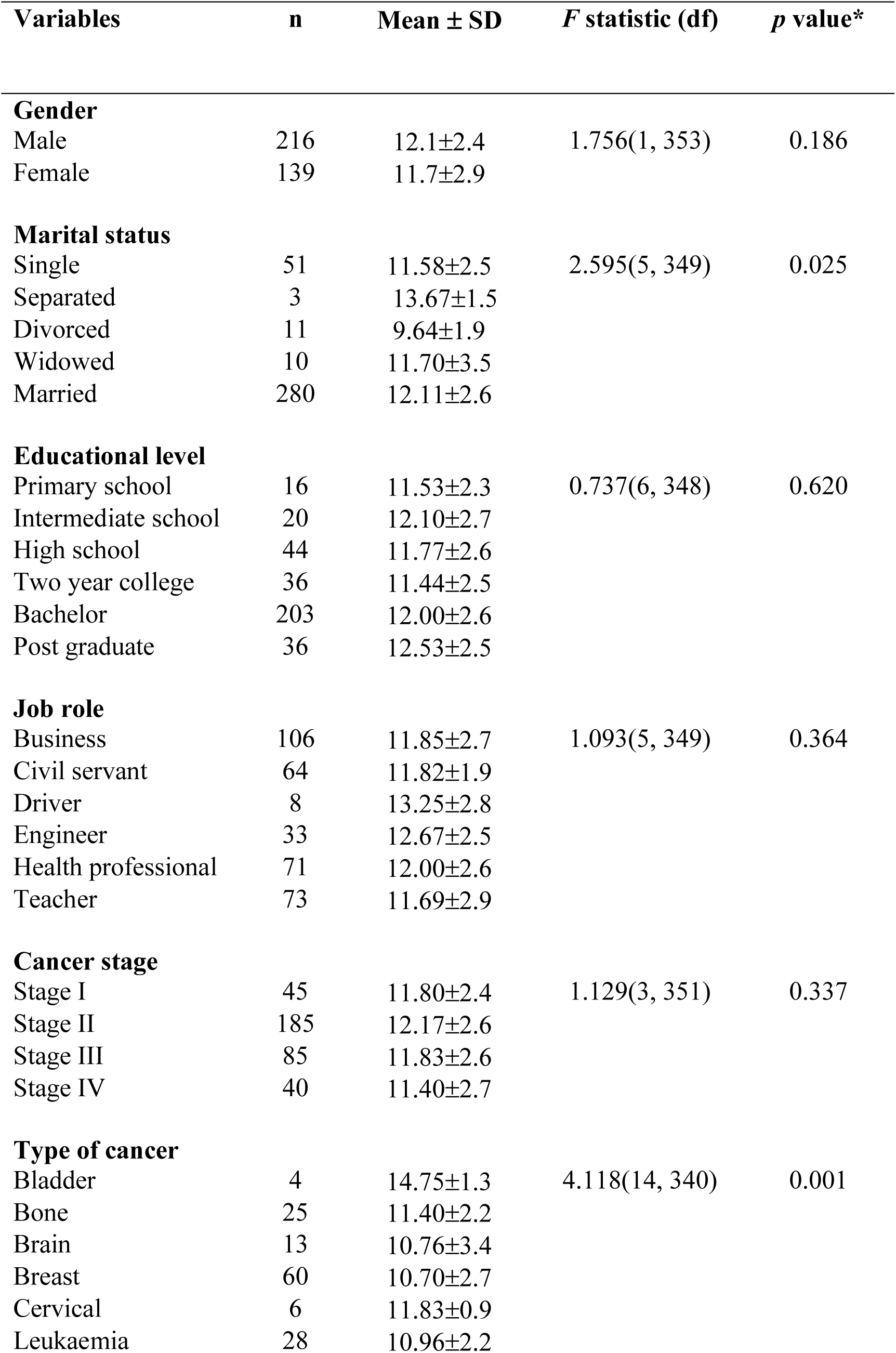
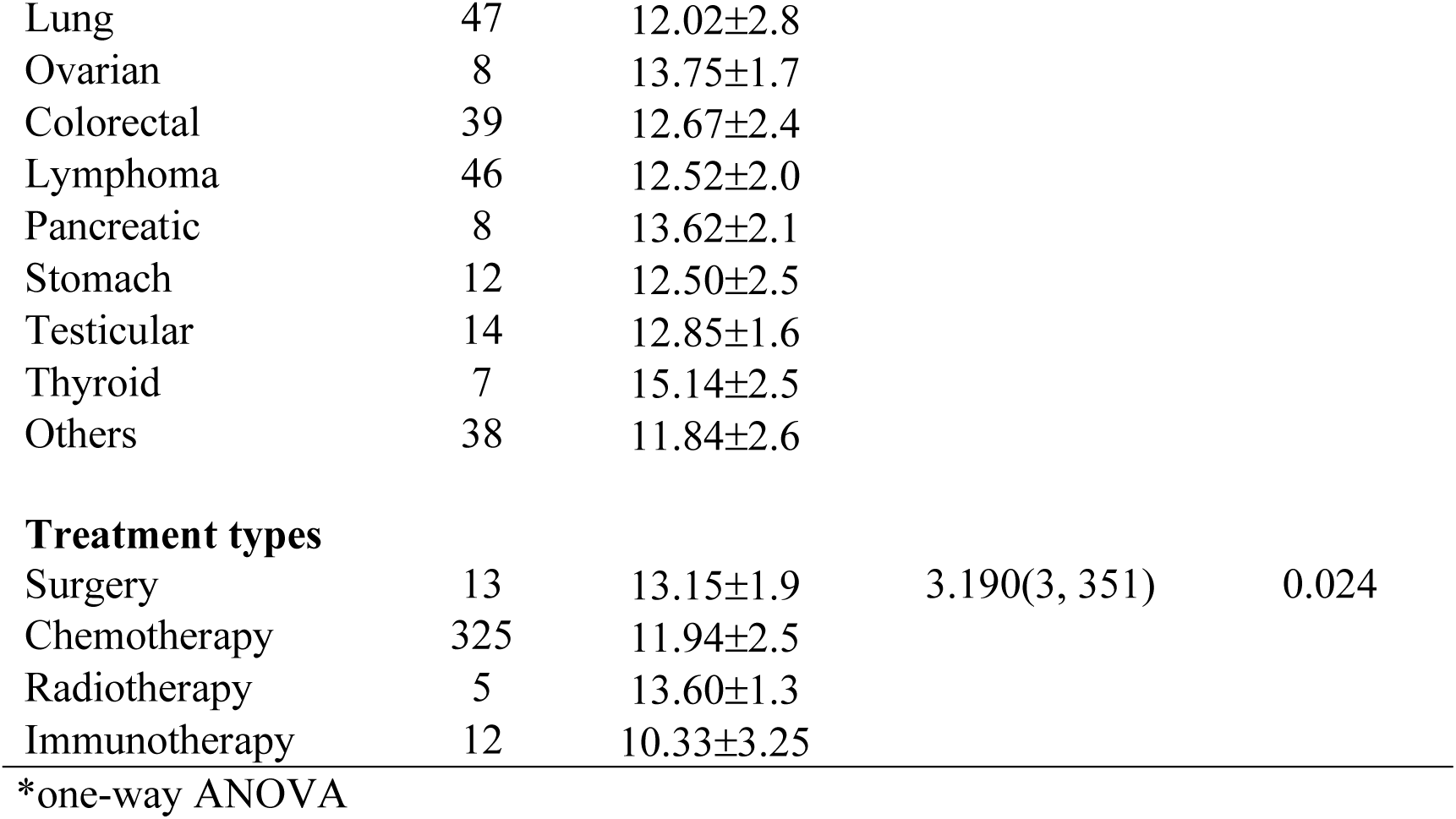
Comparison of means of anxiety disorder among gender, marital status, educational level, job role, cancer stage, type of cancer and treatment types

### Factors contributing to anxiety disorder

Table 6 shows data presentation obtain from simple linear regression which demonstrated that among all the independent variables used to predict anxiety disorder, marital status (β = 0.227, *p* < 0.038), type of cancer (β = 0.131, *p* < 0.001), and treatment types (β = -0.636, *p* < 0.045) have significantly explained or predicted anxiety disorder among workers with cancer. Following a multiple regression analysis (Table 6), it was found that still all the three mentioned variables significantly explained the anxiety disorder. These predictors are types of cancer (β = 0.133, *p* < 0.001) which seems to have higher effect than both marital status (β = 0.197, *p* < 0.036), and treatment types (β = -0.689, *p* < 0.029) on workers’ anxiety disorder. The reported value of the F-statistic (F = 8.117, < 0.001) fits the model data. Standardized Regression coefficients are presented in Table 6 to explain the importance of these predictors on anxiety disorder among workers with cancer. The R_2_ = 0.057 revealed that a combination of this predictor explained 25.5% of variance in anxiety disorder.

**Table 6:**
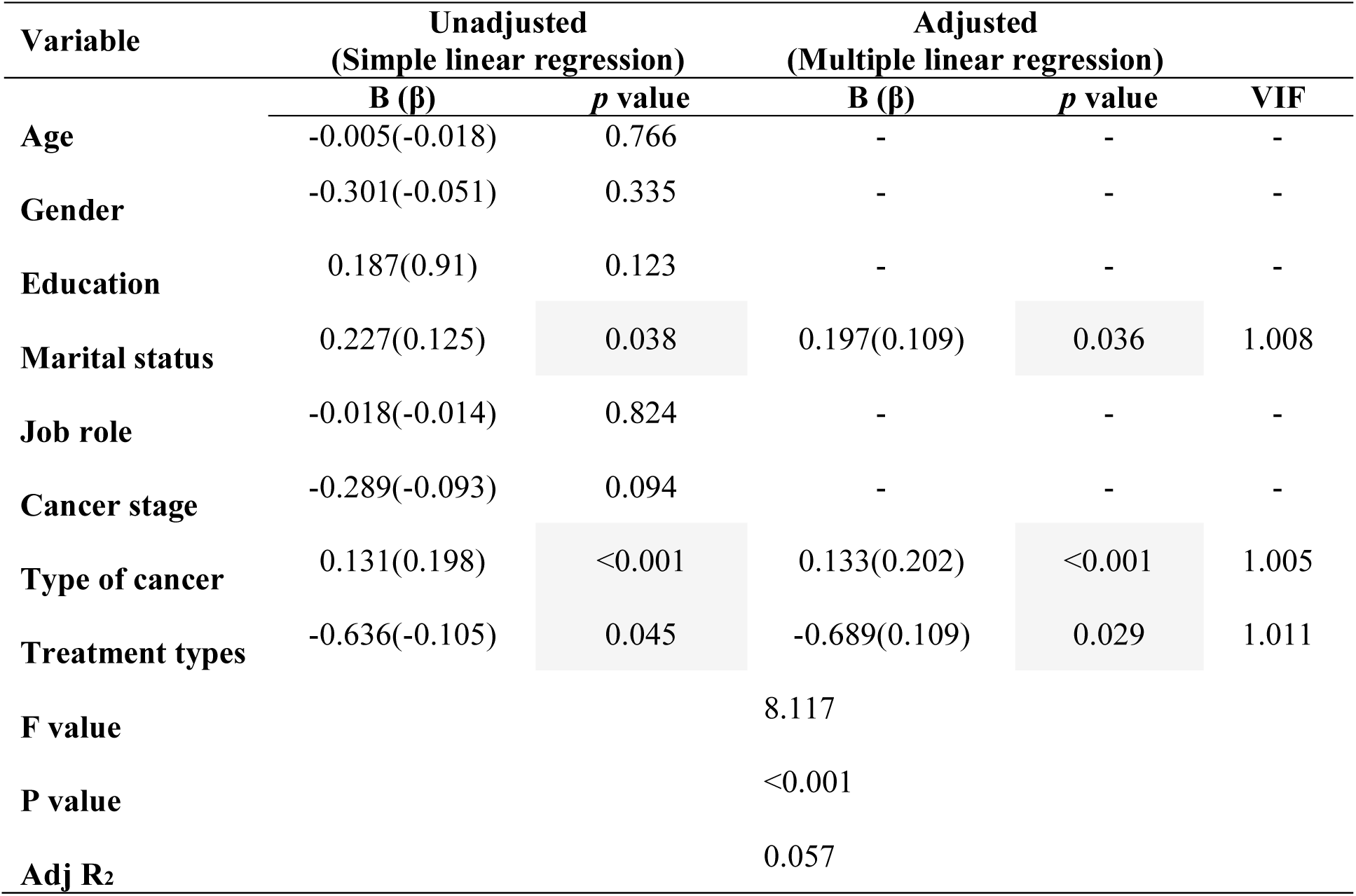
Predictors of anxiety disorder among workers with cancer

### Discussion

This work provides the general prevalence and associated factors of anxiety disorder among 355 workers with cancer who were attending King Husain Cancer Centre (KHCC) in Amman, Jordan. We had a 100% response rate from the participants whose overall meant age was 42.3 years with male at 43.1 years and female aged 41.0 years respectively. More than 70% of the participants were married and about 60% of them had a bachelor degree qualification. Business was the major means of their job role followed by teaching professions. The percentage of these sociodemographic variables are consistent and had no much difference with a study by [18] who explored anxiety and depression among diabetic patients in Amman, Jordan. Our results suggest that majority of the workers with cancer in Jordan were married and averagely, were educated. Analysis of clinical characteristics of workers with cancer showed that majority of the patients were of stage II (52.1%) and stage III cancers (23.9%). And that most of them (91.5%) received chemotherapy drugs as their treatment types while 16.9% were of breast cancer type and most of these patients reported to have had 1-5 months (49.6%) duration of diagnosis. These results suggest high stage tumour is predominant among workers with cancer in Jordan with at least 5 months duration of diagnosis.

However, comparison of these clinical features among workers with cancers is difficult because of the limited studies in this area of research. Hence, this is the first study to report these clinical features in workers with cancer in particularly the Jordanian population. However, cancer staging has been proven to be the most significant prognostic factor for evaluating survival rate[19]. Consistent with other study from Bahrain [20] that demonstrated more cases of patients with cancer were diagnosed at stages II and III and another study carried out in Jordan by [21]who evaluated the epidemiological and survival analysis of Jordanian breast cancer patients from 1997 to 2002. Our findings is contrary to a study conducted in Canada who discovered that about 75% of patients diagnosed with cancer were of stage I and stage II[20] which is unlike in our study that showed majority of the patients fall in stage II and stage III cancer groups. However, stage II is lower in percentage than stage II group in the Canadian population study. This could be due to the fact that hospitals across Jordan implemented a screening programmes for high risks patients who are mostly stage II to be undergoing mammography in combination with regular clinical examinations. Comparison of age based on the mean rank according to the cancer stages showed that patients with stage I and stage II cancer (140.2 vs 168.1) were younger compared with patients with stage III and IV cancer (186.2 vs 249.1) (*p* = 0.001). In contrast, patients with stage II were older than patients with stage I cancer (*p* = 0.001). To best of our knowledge, this is the first population based study that evaluated the impact of age at diagnosis and the clinical features of cancer stages among workers with cancer in Jordan.

Prevalence of anxiety disorder among these workers with cancer was 20.8% accounting for 23.1% male who were seems to have higher prevalence rate than female (17.3%) workers with cancers. Furthermore, prevalent rate of anxiety were noticed mostly among marital status that were separated, widowed, divorce and single. Likewise, high prevalence was observed among professional engineers and business individuals, drivers, civil servant and teachers. Anxiety disorder varies across marital status, types of cancer, and treatment types. The prevalence found in this study is comparable with a study that carried out a systematic review and meta-analysis to demonstrate the prevalence of major and minor depressions as well as anxiety in patients with cancer[22]. Our findings demonstrated high prevalent of anxiety disorder among workers with cancer than can be found in most of the studies[22, 23]. Multilinear regression analysis showed that anxiety is positively associated with marital status (*p* = 0.036), type of cancer (*p* = 0.001), and treatment types (*p* = 0.029) with combination of these predictors explaining 25.5% of variance in anxiety disorder. The prevalence of anxiety in patients with cancer in this study was similar to those of the previous findings that report even high elevated symptoms of anxiety [24, 25]. This could be due to different methodological scale used in measuring the anxiety disorder among the patients. In contrast to our report,[26] reported a higher prevalence of anxiety in breast cancer patients accounting for about 46.8%. Our results also identified that married workers with cancer have less symptoms of anxiety. This finding is similar with recent study conducted in Vietnam that evaluated anxiety among patients with cancer through a hospital based cross-sectional study [27]. Reason for this is because generally, married patients with cancer perceive more social support than unmarried or separated individuals. Anxiety and depression are frequently diagnosed in patients with cancer and are serve the best way of identifying those patients who are in risk of psychological stress [28, 29]. Moreover, anxiety and depression has been linked with poorer physical function [30] and high risk of high mortality in patients with cancer [31].

This study faced some challenges and limitations. Firstly, the sample may not represent the whole population of workers with cancer in Jordan. Even though the Jordanian population is small, but KHCC serve as the largest hospitals where all patients are refereed from all medical facilities in the country. Secondly, the study did not cover some other clinical features due to inaccurate records that were made available to the researcher.

## Conclusion

This study reports the prevalence of anxiety disorder among workers with cancer who were attending King Husain Cancer Centre (KHCC) in Amman, Jordan. Our findings demonstrated higher prevalent rate of anxiety among these patients. Marital status, type of cancer and treatment types was the major predictors of anxiety among workers with cancer. However, other unknown factors might be the cause for this high prevalent of anxiety which need to be further investigated. This high level of anxiety and distress experienced by workers with cancer is a serious public health issues that required government attention for effective work delivery among workers with cancer. Thus, workers with cancer need to be monitor and screened for anxiety disorder especially when register for clinical check-up as this will help in drastically reducing the distress they might have faced during the diagnostic period.

## Data Availability

All data are analysed and reported in this manuscript.

## Acknowledgments

The authors extend their special thanks to the management and staff of King Hussain Medical Centre in Amman, Jordan for allowing us to embarked on this study using their attending patients. We are also indebted to University Ethics Committee for Research involving Human Subjects, Faculty of Medicine and Health Sciences (FMHS), University Putra Malaysia (UPM) for approving the study.

